# Cross-neutralizing activity against SARS-CoV-2 variants in COVID-19 patients: Comparison of four waves of the pandemic in Japan

**DOI:** 10.1101/2021.06.10.21258682

**Authors:** Koichi Furukawa, Lidya Handayani Tjan, Silvia Sutandhio, Yukiya Kurahashi, Sachiyo Iwata, Yoshiki Tohma, Shigeru Sano, Sachiko Nakamura, Mitsuhiro Nishimura, Jun Arii, Tatsunori Kiriu, Masatsugu Yamamoto, Tatsuya Nagano, Yoshihiro Nishimura, Yasuko Mori

**Affiliations:** Division of Clinical Virology, Center for Infectious Disease, Kobe University Graduate School of Medicine, Kobe, Hyogo 650-0017, Japan; Division of Cardiovascular Medicine, Hyogo Prefectural Kakogawa Medical Center, Kakogawa 675-0003, Japan; Acute Care Medical Center, Hyogo Prefectural Kakogawa Medical Center, Kakogawa 675-0003, Japan; Division of General Internal Medicine, Hyogo Prefectural Kakogawa Medical Center, Kakogawa 675-0003, Japan; Division of Respiratory Medicine, Department of Internal Medicine, Kobe University Graduate School of Medicine, Kobe, Hyogo 650-0017, Japan

**Keywords:** SARS-CoV-2, COVID-19, variant, reinfection, neutralizing activity

## Abstract

In March 2021, Japan is facing a 4th wave of SARS-CoV-2 infection. To prevent further spread of infection, sera cross-neutralizing activity of patients previously infected with conventional SARS-CoV-2 against novel variants is important but is not firmly established. We investigated the neutralizing potency of 81 COVID-19 patients’ sera from 4 waves of pandemic against SARS-CoV-2 variants using their authentic viruses. Most sera had neutralizing activity against all variants, showing similar activity against B.1.1.7 and D614G, but lower activity especially against B.1.351. In the 4th wave, sera-neutralizing activity against B.1.1.7 was significantly higher than that against any other variants, including D614G. The cross-neutralizing activity of convalescent sera was effective against all variants but was potentially weaker for B.1.351.

## BACKGROUND

The coronavirus disease 2019 (COVID-19) pandemic declared by the World Health Organization (WHO) in March 2020 continues to affect all countries around the world. In efforts to control the pandemic, several vaccine platforms have been developed based on the original severe acute respiratory syndrome coronavirus 2 (SARS-CoV-2) (Wuhan-1) as the template, and these vaccines have been shown to be effective for reducing the COVID-19 outbreak [1-3].

However, the evolution of SARS-CoV-2 has continued since its initial emergence. By the beginning of April 2020, a variant bearing a D614G mutation with evidence of increased infectivity had become dominant [4]. The SARS-CoV-2 variant B.1.1.7, first detected in Kent and Great London in September 2020, has now spread to many countries worldwide with evidence indicating an increased mortality rate [5, 6]. In addition to D614G and several mutations in other areas of the genome, B.1.1.7 bears eight mutations in the spike gene including deletions in the N-terminal domain (ΔH69/ΔV70, Δ144) and amino acid substitutions in the receptor binding domain (N501Y) [7, 8].

The SARS-CoV-2 variant B.1.351 was first detected in specimens collected from South Africa in October 2020, and it has rapidly become the predominant variant circulating throughout South Africa [9]. Among the nine mutations in the spike gene in this variant, there are three biologically important mutations: K417N, E484K, and N501Y [7]. Importantly, there is growing evidence that the B.1.351 variant has the ability to escape from the neutralizing antibody elicited against the original SARS-CoV-2 infection and currently available vaccines [7, 10-12].

The SARS-CoV-2 variant P.1, which was first detected in Japan in early January 2021 from four individuals with a history of traveling to Brazil, had become the predominant variant circulating in Brazil by January 2021 [13]. It bears 12 mutations in the spike gene, including K417T, E484K, and N501Y [14], which are the same three amino acid substitutions as those found in B.1.351. Interestingly, variant P.1 showed less resistance to a neutralizing antibody induced by natural infection or vaccination compared to a similar variant, B.1.351 [15].

The emergence of these variants poses a tremendous challenge to the control of the SARS-CoV-2 pandemic. In addition, the B.1.351 and P.1 variants carry the E484K mutation that is responsible for evasion from the monoclonal antibody against original SARS-CoV-2, further compromising the currently available therapy against this virus [16].

As of May 2021, Japan has experienced four waves of the COVID-19 pandemic, beginning in April 2020; the number of total confirmed cases is over 690,000 and there have been more than 11,000 deaths due to COVID-19 in Japan alone [17]. The growth rate of the number of infected individuals in the 4th wave is much faster than those of the 1st to 3rd waves so far, and there is concern about the possibility of a collapse of the healthcare system. SARS-CoV-2 genome surveillance has revealed that D614G_KR and its lineages were the predominating circulating viruses responsible for the 1st to 3rd waves of the pandemic in Japan, but the introduction of the R1 and B.1.1.7 variants in late 2020 has replaced the previously existing strains and may be responsible for the 4th wave [18]. The B.1.351 and P.1 variants have also been detected in Japan, although no trend toward an increasing dominance of these variants has been observed thus far [19].

It is not yet known to what extent the serum of patients previously infected with original SARS-CoV-2 might confer protection against these rapidly emerging variants. In this study, we investigated the neutralizing potency of serum from patients infected during the 1st to 4th waves of the pandemic against SARS-CoV-2 variants D614G, B.1.1.7, B.1.351, and P1, using authentic virus. This research is imperative to understand whether individuals who have recovered from COVID-19 could be protected from reinfection by newly emerging variants. This research might also help predict the potency of using plasma from individuals who recovered from the conventional type or any variants of SARS-CoV-2 as a donor if convalescent plasma therapy can be used for COVID-19 patients infected by the other variants.

## METHODS

### Diagnosis of COVID-19

COVID-19 diagnoses were based on the polymerase chain reaction (PCR) detection of the SARS-CoV-2 genome in nasopharyngeal swab samples. Disease severity was defined as follows: Symptomatic COVID-19 cases without evidence of pneumonia or hypoxia were classified as mild. Cases in patients with clinical signs of pneumonia were classified as moderate (oxygen saturation as measured by pulse oximetry, ≥90% with room air) or as severe (respirations >30/min, severe respiratory distress, or oxygen saturation <90% with room air). Patients who needed mechanical ventilation were classified as critical.

### Definitions of the waves of the COVID-19 pandemic in Japan

The period from the 1st wave to the 4th wave of the COVID-19 pandemic was defined based on the change in the number of infected people on a single day in Japan. The 1st wave was from March 1st to the end of June 2020; the 2nd wave was from July 1st to the end of October 2020; the 3rd wave was from November 1st 2020 to the end of February 2021, and the 4th wave was the period beginning March 1st 2021 [17].

### Participant recruitment

From March 2020 to May 2021, blood samples were collected from patients who became infected with SARS-CoV-2 and were hospitalized at Hyogo Prefectural Kakogawa Medical Center (Hyogo, Japan). We selected serum of convalescent patients with different disease severities who were already confirmed to have neutralizing activity against the SARS-CoV-2. In May 2020, the serum of 24 healthy individuals were collected and confirmed to have no antibody against SARS-CoV-2; these sera were used as the negative control group [20]. This study was a retrospective observational investigation and was carried out after written consent was obtained from the subjects or by the opt-out method when it was difficult to get written consent due to the disease severity. No statistical methods were used to predetermine the sample size.

### Measurement of neutralizing activity against SARS-CoV-2

Neutralization was performed as described [21]. Briefly, the neutralizing activity of each serum sample was evaluated by a neutralization assay against each living SARS-CoV-2 variant (D614G, B.1.1.7, P.1, or B.1.351) in a biosafety level 3 laboratory. Vero E6 (TMPRSS2) cells were used [22]. The neutralizing antibody titer was determined as the highest serum dilution that did not show any cytopathic effects.

### Preparation of SARS-CoV-2 variants

We used the SARS-CoV-2 Biken-2 (B2) strain with a D614G mutation as a conventional variant. It was provided by the Research Foundation for Microbial Diseases of Osaka University (BIKEN). The three variants B.1.1.7, P.1, and B.1.351 were isolated and provided by the National Institute of Infectious Disease, Japan.

### Statistical analysis

GraphPad Prism software (ver. 8.4.3) was used for the statistical analysis and preparation of figures. The Friedman test was used to compare the neutralizing antibody titer among the four variants. The Kruskal-Wallis test was used to compare the neutralizing antibody titer among different disease severity groups. Results were considered significant at a p-value <0.05.

### Ethical approval

This study was approved by the ethical committees of Kobe University Graduate School of Medicine (approval code: B200200) and Hyogo Prefectural Kakogawa Medical Center.

## RESULTS

### Patient characteristics

We examined a total of 81 sera of patients with different disease severities who were already confirmed to have neutralizing activity against B2 strain, which is a D614G variant. The median number of days between the onset of symptoms and the collection of serum samples (days post-onset, dpo) was 26 days. Overall, 62% of the patients were male, 38% were female, and the median age was 64 years. The asymptomatic/mildly infected group was comprised of 25 patients, 19 patients were moderate/severe, and the remaining 37 patients were in the critical infection group. The most common medical histories were hypertension and diabetes, in 28.4% of the patients each.

Eleven patients had received antiviral treatment with favipiravir or lopinavir (both for six patients and favipiravir for five patients), and 42 patients received steroid treatment. A comparison of the four waves revealed that the 2nd wave (with 20 patients) contained only one critical patient, whereas all 20 patients in the 4th wave were critical and were mostly (75%) male. In addition, antiviral treatment was mainly prescribed for the patients in the 1st wave, whereas steroids were mainly used in the 2nd wave onward.

### Neutralizing activity against all variants in all patients

Most of the 81 sera had neutralizing activity against the four variants, although the activity values varied (Fig. 1). The mean neutralizing antibody titer for the D614G variant was 80, and that for the B.1.1.7 variant was 111. The neutralizing titer of B.1.1.7 seemed to be higher than that of D614G, but the difference was not significant. In contrast, the mean neutralizing antibody titer against P.1 was 44 and that against or B.1.351 was 21, and each of these values was lower than that for D614G, especially in B.1.351 (3.8x, p<0.0001).

**Fig. 1.**
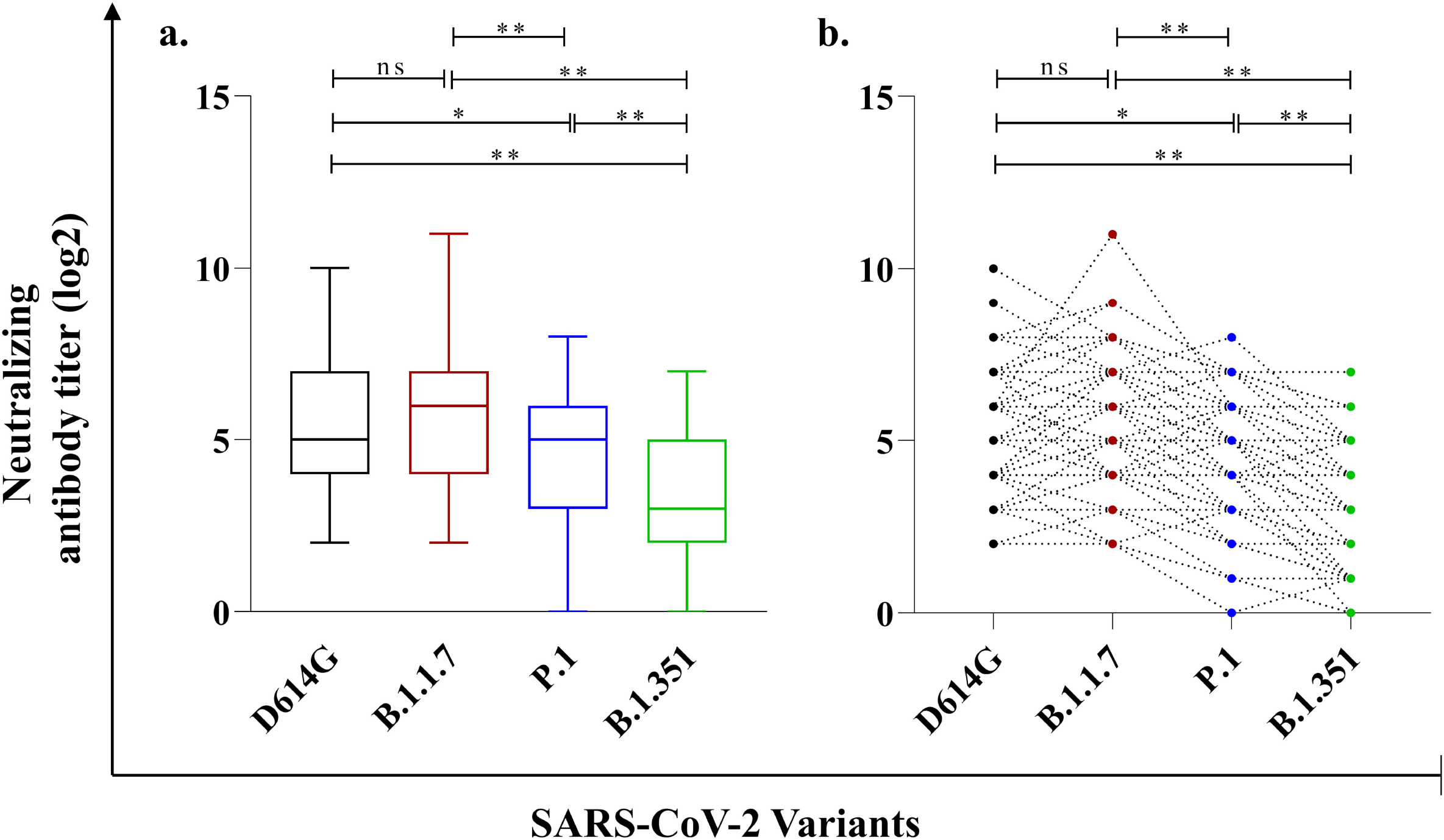
Neutralization activity against SARS-CoV-2 variants. Sera of 81 patients who had recovered from COVID-19 were tested for neutralizing activity against the SARS-CoV-2 variants D614G, B.1.1.7, P.1, and B.1.351. The neutralizing antibody titer is represented by the logarithmic scale of the highest serum dilution that did not show any cytopathic effects. **a:** Box plot of the neutralizing antibody titers with the minimum, first quartile, median, third quartile, and maximum values. **b:** Changes in the antibody titer for each patient. The titer of the same patient is connected by a line. The Friedman test was used, and two-tailed p-values were calculated. *p<0.05, **p<0.01.

### Neutralizing activity against all variants in each wave

From the 1st wave to the 3rd wave, the neutralizing activity against B.1.1.7 variant was similar or slightly low compared to that against D614G, whereas it was higher in the 4th wave (increased 4x, p = 0.0009). In addition, the neutralizing activity against B.1.1.7 was also higher than that against P.1 or B.1.351 variant in the 4th wave. In all waves, the neutralizing activity against B.1.351 variant was lower than those against the other three variants (Fig. 2).

**Fig. 2.**
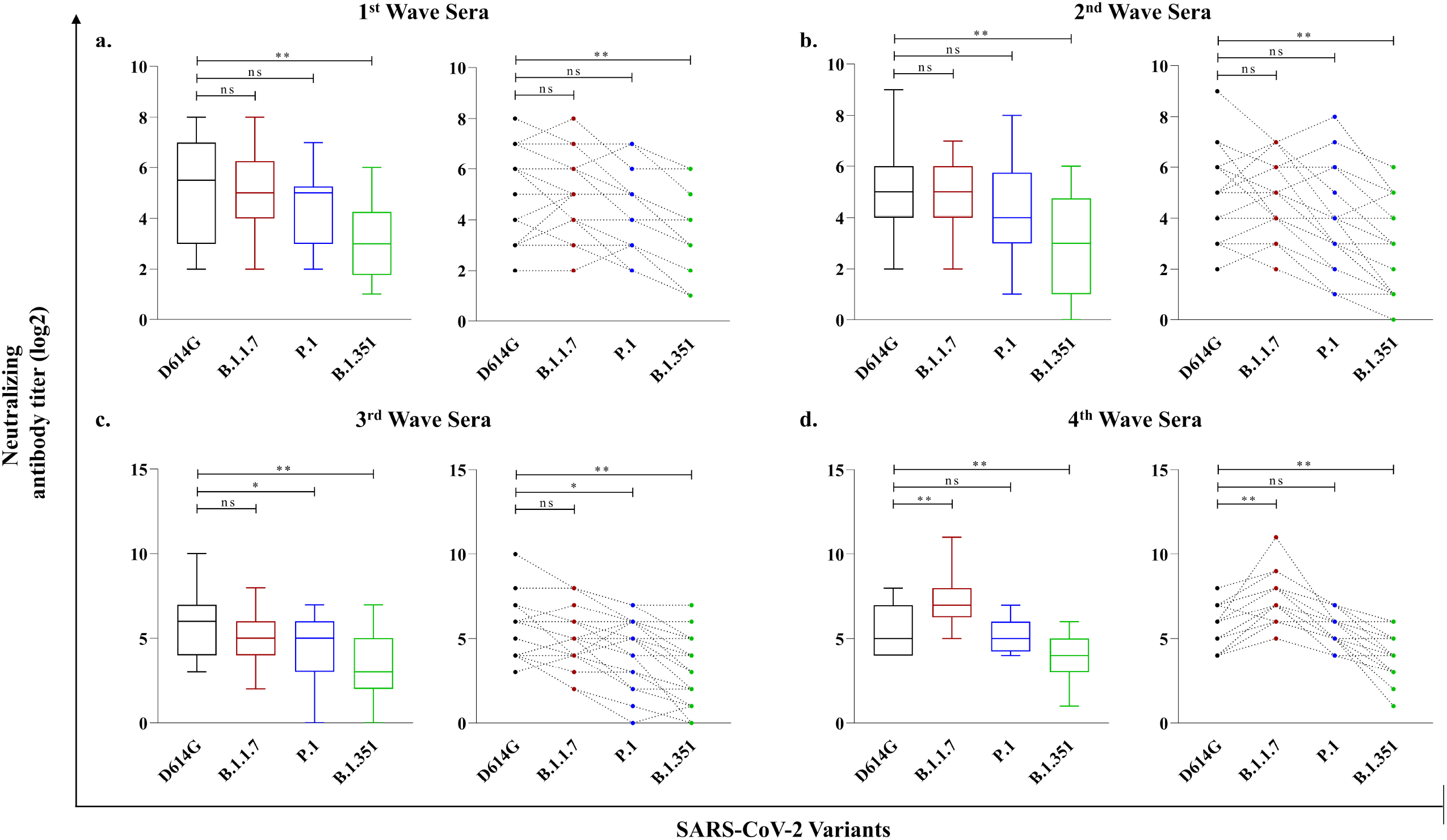
The neutralizing activity against all variants in each wave. The neutralizing antibody titers of sera against D614G, B.1.1.7, P.1, and B.1.351 were compared in the 1st wave (from March 1st to June 2020, **a**), 2nd wave (from July 1st to October 2020, **b**), 3rd wave (from November 1st 2020 to February 2021, **c**) and 4th wave (after March 1st 2021, **d**). The Friedman test was used, and two-tailed p-values were calculated. *p<0.05, **p<0.01.

### Neutralizing activity against each variant by severity

The sera of all of the COVID-19 patients showed neutralizing activity against the D614G and B.1.1.7 regardless of the severity of the patients’ symptoms. A significantly lower neutralizing titer against D614G, B.1.1.7, P.1, or B.1.351 was observed in the serum of the asymptomatic/mild COVID-19 patients compared to that of the critical patients (four-to ninefold lower, p<0.0001) (Fig. 3a–d).

**Fig. 3.**
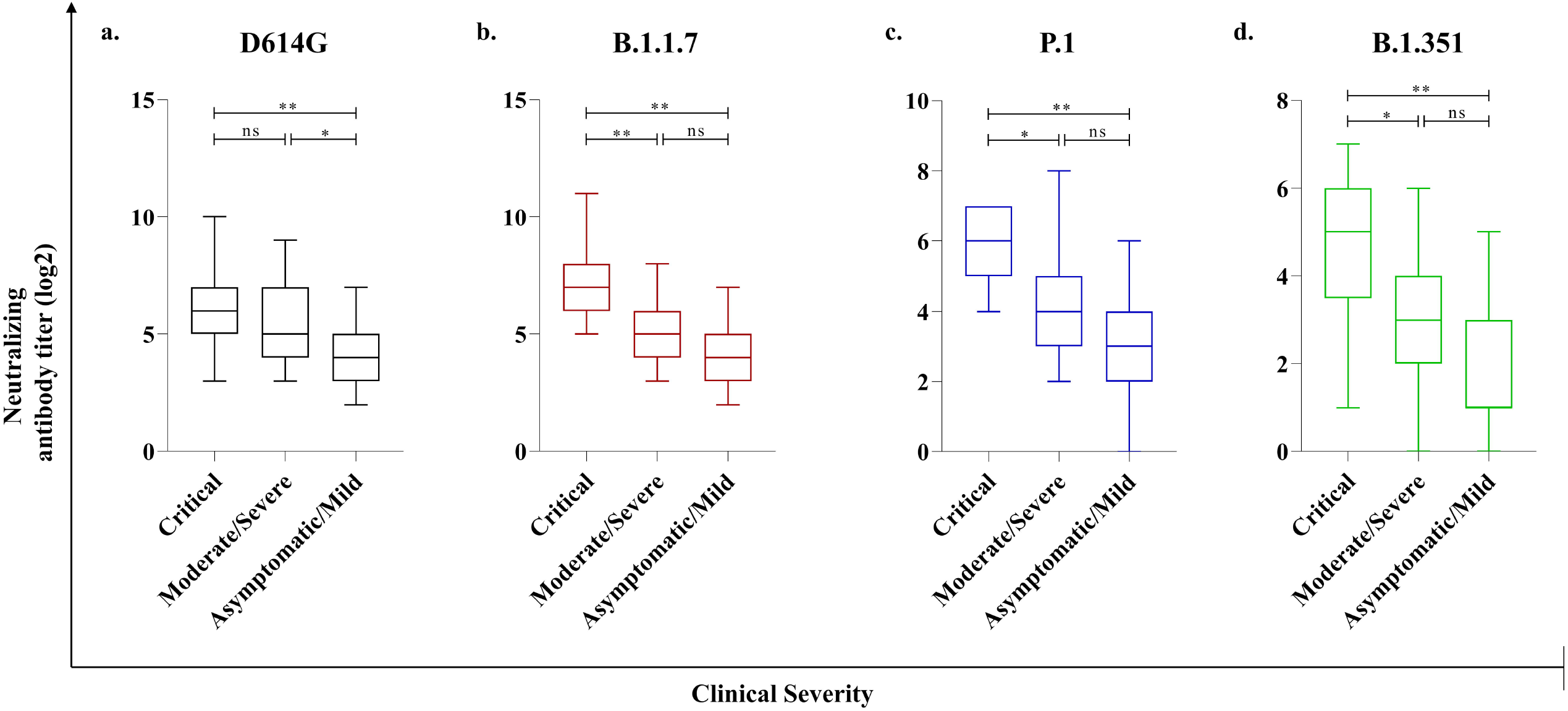
The neutralizing activity against each variant by disease severity. The neutralizing antibody titer against **(a)** D614G, **(b)** B.1.1.7, **(c)** P.1 and **(d)** B.1.351 in patient’s sera with different severity groups. The Kruskal-Wallis test was used, and two-tailed p-values were calculated. *p<0.05, **p<0.01.

Interestingly, almost all of the sera from the asymptomatic/mild infected group, with the exception of three cases, had neutralizing activity against all tested variants. Three asymptomatic/mild cases and one case in the severe-infection group with low neutralizing activity against D614G (titer 8 or 16) did not show any neutralizing activity against P.1 or B.1.351 (Fig. 3c,d).

## DISCUSSION

In Japan, the 4th wave of SARS-CoV-2 arrived in March 2021, and the presence of the variant B.1.1.7 has increased in this wave. It is suspected that the conventional D614G variant has already been almost completely replaced by B.1.1.7. In addition, P.1 and B.1.351 have also been identified in Japan, and there is thus a possibility of a further spread of infection in the future. Given the recent emergence of the B.1.1.7, P.1, and B.1.351 variants, the cross-neutralization of these variants by previous pandemic sera remains to be clarified. To predict and help prevent the further spread of SARS-CoV-2 infection, it is necessary to determine whether the neutralizing activity in COVID-19 patients infected with the D614G have similar activity against the newly emerging variants.

In the present study, regardless of the patients’ infection time (wave) and disease severity, most of their sera had neutralizing activity against the four variants (D614G, B.1.1.7, P.1, and B.1.351) although the neutralizing activity values varied. Some individuals that showed high neutralizing activity against D614G and B.1.1.7, also had the high activity against P.1 and B.1.351, indicating that individuals infected with D614G or B.1.1.7 also could have the neutralizing antibody against P.1 and B.1.351.

Although we observed no significant difference between the neutralizing activity of sera against B.1.1.7 and D614G in all patients, the values of neutralizing activity against P.1 and B.1.351 were lower than that against D614G, and the neutralizing activity against B.1.351 in particular was much lower. This means that the neutralizing activities of sera from previously infected patients was also seen against the B.1.1.7 but was potentially weaker against the P.1 and B.1.351. As one of the potential explanations for this finding, we note that N501Y substitution (which is common among these three variants) may enhance the binding to ACE2, but its antigenic effects are limited and it may little affects the neutralizing activity of the antibodies [23, 24]. However, E484K mutation which is found both in P.1 and B.1.351, but not in either D614G and B.1.1.7, has been reported to affect the binding of serum polyclonal neutralizing antibodies [16].

On the other hand, because P.1 and B.1.351 have similar mutations in their RBD (including E484K, K417T/N, and N501Y), it might be thought that the neutralization of both variants would be affected similarly. However, our present analyses demonstrated that while some sera of individuals showed similar or high neutralizing activity against P.1 compared to those against D614G, the activity against B.1.351 was consistently lower than that against D614G, indicating that B.1.351 might avoid the neutralization more effectively by means other than mutations of the RBD, such as the amino acid deletions (242-244 del) and substitutions (D80A, R246I) in the N terminal domain (NTD) [7, 11, 25].

Interestingly, although we observed that the neutralizing activity against the B.1.1.7 seemed to be similar to or slightly lower than that against D614G from the 1st to 3rd waves in Japan, its activity against B.1.1.7 was higher than that against D614G, P.1, and B.1.351 in the 4th wave, indicating an epidemic of B.1.1.7. In particular, the neutralizing activities against P.1 and B.1.351 were significantly lower than that for B.1.1.7. Regarding this result, some other groups have also reported that antibodies elicited by B.1.1.7 infection exhibited significantly reduced recognition and neutralization of parental (Wuhan) strain or B.1.351 compared to B.1.1.7 [26, 27]. Our result may suggest that the mutations in B.1.1.7 could cause the conformational change of its spike protein, which affects the immune recognition for D614G.

The correlation between serum neutralization activity against D614G and clinical severity has been described [28-31], and our present findings revealed a similar correlation for three other variants. Even among the asymptomatic/mild patients, all had neutralizing activity against B.1.1.7 and most also had neutralizing activity against P.1 and B.1.351.

Our results suggest that natural infection with each SARS-CoV-2 variant prompts the body to make antibodies that recognize the infecting strain most robustly, with various degrees of cross-recognition of other strains. The efficacy of convalescent plasma therapy remains controversial, but it may be considered to use the convalescent sera induced by conventional strain for high risk patients infected with B.1.1.7 or P.1 [32-34]. Individuals recovered from the infection of 4^th^ wave may not completely

protect against reinfection with the other SARS□CoV□2 variants in the future, especially in asymptomatic or mild cases which have low neutralizing activity. Our findings may indicate that the cross-neutralization could work to protect against the induction of severe symptoms when an individual is reinfected by new variants. Further studies are required to address this and many other questions about the variants that continue to arise.

## Data Availability

The authors declare no conflicts of interest with respect to this.

## Acknowledgments

We thank Kazuro Sugimura MD, PhD (Kobe University and Hyogo Prefectural Hospital Agency) for his full support to promote this study. We express our sincere gratitude for cooperation and participation of staffs of Hyogo Prefectural Kakogawa Medical Center. We thank Research Foundation for Microbial Diseases of Osaka University (BIKEN), Osaka University for providing SARS-CoV-2 B2 strain. We thank the National Institute of Infectious Disease Japan for providing SARS-CoV-2 B.1.1.7, P.1, and B.1.351 variants.

